# Influence of Incorrect Documentation of Initial Cardiac Rhythm in Survivors of Out of Hospital Cardiac Arrest

**DOI:** 10.1101/2024.04.23.24306258

**Authors:** Sant Kumar, Max Hockstein, Alexander I. Papolos, Mithun Devraj, Nauman Khalid, Michael J. Lipinski, Benjamin B. Kenigsberg

**Affiliations:** MedStar Georgetown University Hospital, Washington, DC; Department of Critical Care, MedStar Washington Hospital Center, Washington, DC; Divison of Cardiology, MedStar Washington Hospital Center, Washington, DC; The George Washington University Heart & Vascular Institute, Washington, DC; St. Francis Medical Center, Monroe, LA; Cardiovascular Associates of Charlottesville, Charlottesville, VA

**Author notes:** **Corresponding Author**: Benjamin B. Kenigsberg, MD FACC, Department of Critical Care and Division of Cardiology MedStar Washington Hospital Center, 110 Irving Street NW, Washington, DC 20010.

**Keywords:** Cardiac arrest, Shockable rhythm, Non-shockable rhythm, Catheterization

## Abstract

**Background:** In-hospital management of out-of-hospital cardiac arrest (OHCA) is heavily influenced by the initial rhythm (shockable vs. non-shockable). The prevalence of discordance between initial rhythm determination reported by emergency medical services (EMS) versus hospital teams is not well described. It is unclear whether such documentation discrepancies in OHCA influence inpatient clinical care including subsequent left heart catheterization (LHC). We hypothesized that discordance between EMS and hospital team documentation of OHCA initial rhythm was common and associated with differences in LHC frequency.

**Methods:** This was a retrospective, single-center study. OHCA patients from the Cardiac Arrest Registry to Enhance Survival (CARES) hospital database were linked by demographic and arrest history to OHCA patients identified by inpatient hospital billing codes from 2009 to 2017. Patients who expired within 24 hours of hospital presentation were excluded. Hospital documentation of OHCA initial rhythm and occurrence of LHC were manually reviewed. The relationship between EMS versus hospital team documentation of OHCA initial rhythm and occurrence of LHC were assessed by relative risk ratios with 95% confidence intervals.

**Results:** Out of 164 patients for analysis, 140 (85.4%) had concordant EMS and hospital documentation of OHCA initial rhythm. For OHCA with an EMS-documented shockable rhythm, the relative risk of LHC when hospital-documented concordant shockable vs. discordant non-shockable rhythm was 2.12 (95% RR CI: 0.76-5.93). For OHCA with an EMS-documented non-shockable rhythm, the relative risk of LHC when hospital-documented concordant non-shockable vs discordant shockable rhythm was 0.19 (95% CI: 0.05-0.69).

**Conclusions:** In patients with OHCA, discrepancy between EMS and hospital team documentation of the initial arrest rhythm is prevalent. This discrepancy may influence the incidence of LHC. Further research is needed to understand the clinical impact of discrepancies in rhythm communication between EMS and hospital teams.

## INTRODUCTION

Initial cardiac rhythm strongly influences outcome after out of hospital cardiac arrest (OHCA). OHCA due to asystole or pulseless electrical activity, also referred to as non-shockable rhythms, is associated with decreased survival. This is attributed, in part, to more frequent non-cardiac etiologies to the arrest.[1],[2] Alternatively, OHCA due to ventricular tachycardia or ventricular fibrillation, also referred to as shockable rhythms given the required intervention with defibrillation, is associated with higher survival and a greater incidence of intervenable cardiac arrest etiologies such as an acute coronary ischemia.[3] As a result, many patients who experience OHCA, particularly from shockable rhythms, are referred for coronary angiography.[3]

However, OHCA events occur in an unpredictable manner and in unpredictable locations, and thus, direct assessment of the initial cardiac rhythm may be unavailable to the hospital-based medical team.[4] Emergency medical services (EMS) are typically the first medical responders to an OHCA and have directly assessed the initial cardiac rhythm, occasionally from an automated external defibrillator (AED). The accuracy of communication of OHCA initial rhythm from EMS to hospital teams is unknown. Additionally, it is also unknown whether discrepancies in OHCA initial rhythm documentation between EMS and hospital records are associated with different frequencies of coronary angiography.

The aim of this study was to assess the frequency of concordance and discordance between EMS and hospital team documentation of initial cardiac rhythm in OHCA. We additionally sought to characterize associated clinical management differences in the disparate groups.

## METHODS

### Source of Data and Study Patients

This was a retrospective, single-center study in which we abstracted data from our electronic health record using billing codes for cardiac arrest from March 2009 to March 2017. Abstracted data included demographics, initial OHCA rhythm from admission documentation, and incidence of invasive cardiac procedures (including left heart catheterization, right heart catheterization and percutaneous coronary intervention). Right heart catheterization, or pulmonary artery catheter placement, was to assess cardiogenic shock. Patients were then associated with their entries in the Cardiac Arrest Registry to Enhance Survival (CARES) registry based on age, date of presentation, and gender. The CARES registry is an online-based data management system in which participating 911 centers, first responders, emergency medical services (EMS) and hospitals enter local data to develop standard outcome measures for OHCA and to improve quality of care. The CARES database lists six options for the initial rhythm recorded by paramedics: “ventricular fibrillation,” “ventricular tachycardia,” “unknown shockable rhythm,” “asystole,” “idioventricular/PEA,” and “unknown unshockable rhythm.”[5] This determination by the paramedics was utilized to identify the EMS documented binary shockable (ventricular fibrillation, ventricular tachycardia and unknown shockable rhythm) and non-shockable (asystole, idioventricular/PEA and unknown unshockable rhythm) rhythm cohorts. When a cross-referenced record was not clear based on basic demographic data alone, available OHCA arrest details including location of the arrest were manually reviewed based on date of OHCA. To ensure consistent admission documentation was available, patients who expired within 24 hours of hospital presentation were excluded from analysis (**Figure 1**). When the hospital team recorded that patients had both shockable and non-shockable rhythms, we classified the initial rhythm as shockable. Additionally, the initial hospital ECG of each patient was manually reviewed by a study physician for evidence of ischemia; however, the initial rhythm strip from the arrest was not available for retrospective review and rhythm verification.

**Figure 1.**
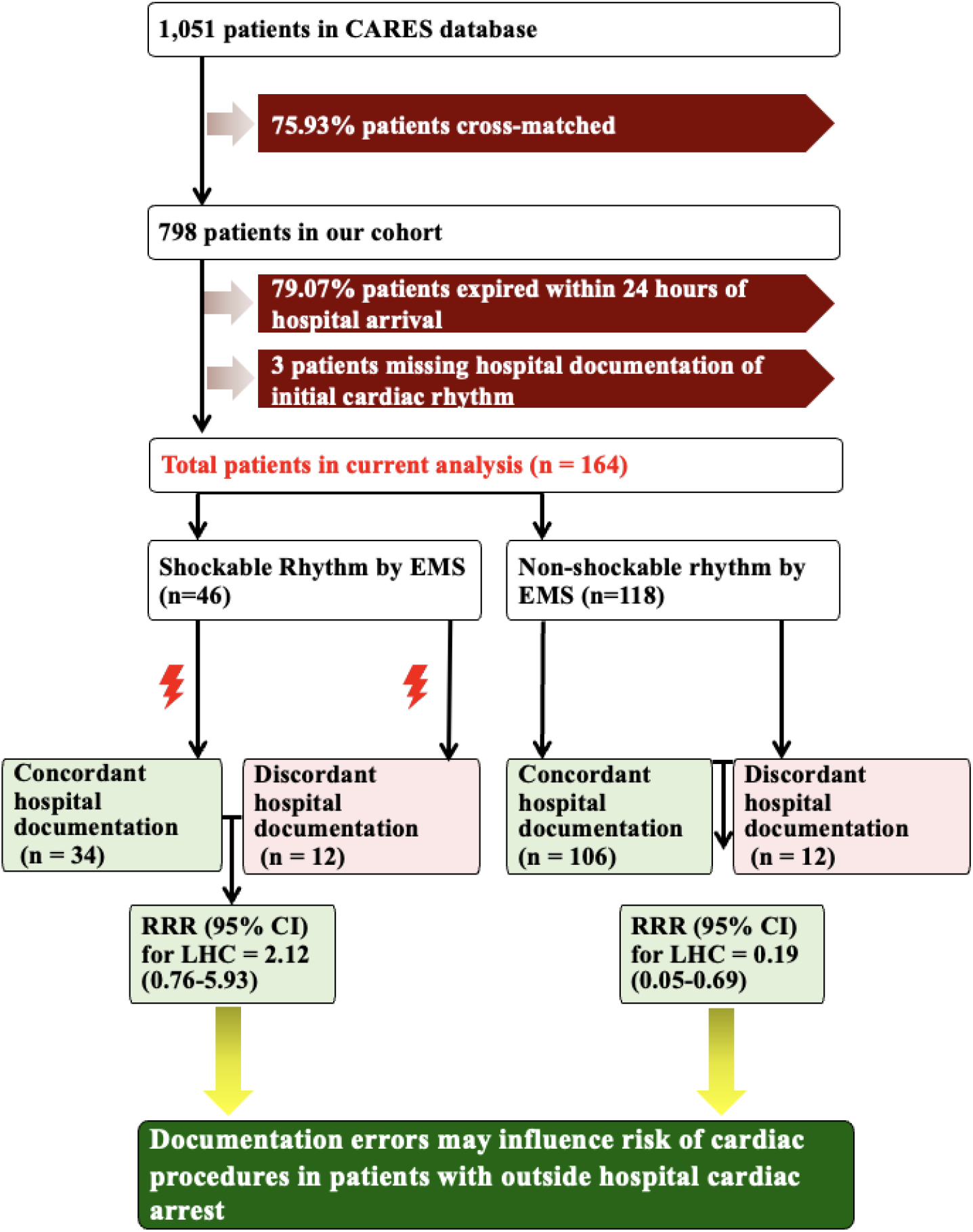
Assembly of cohort and impact of incorrect documentation of initial cardiac rhythm, stratified by initially documented EMS rhythm Flow chart displaying assembly of cohort and potential impact of discordance between EMS and hospital documentation of initial cardiac rhythm, stratified by EMS documentation of initial cardiac rhythm. Abbreviations: EMS = emergency medical serves; RRR = relative risk ratio; CI = confidence interval; LHC = Left heart catheterization

While our institutional practice has evolved over time with developing data, we generally adhere to the current Society for Cardiovascular Angiography and Interventions (SCAI) recommendations for managing patients with OHCA.[6] Specifically, our typical approach involves immediate LHC with coronary angiography and primary percutaneous coronary intervention (PCI) as indicated for patients with either intact neurological function or comatose patients with favorable features for good neurological recovery, who have ST-segment elevation myocardial infarction (STEMI) on their initial ECG or evident ischemia mediated cardiogenic shock after the return of spontaneous circulation (ROSC). We consider LHC urgently for patients with shockable initial OHCA rhythms and/or non-ST-segment elevation myocardial infarctions (NSTEMI), although over time have generally prioritized awaiting neurological recovery before proceeding with LHC. We consider LHC on a case-by-case basis in patients with OHCA and non-shockable rhythms, considering post ROSC ECG, cardiac biomarkers, clinical status and medical histories.[6] The study protocol was approved by the IRB at MedStar Washington Hospital Center.

### Outcome Definitions

#### Concordant Documentation

If hospital documentation of the initial cardiac rhythm in patients with OHCA matched the rhythm documented by EMS, we defined this as “concordant.” For example, if a patient with OHCA was noted to have a shockable rhythm (ventricular tachycardia, ventricular fibrillation or unknown shockable rhythm) by EMS, and the hospital team noted the initial out of hospital arrest rhythm to be shockable (e.g. ventricular tachycardia or ventricular fibrillation) in its documentation as well, then we defined this as concordant documentation (**Figure 1**).

#### Discordant Documentation

If hospital documentation of the initial cardiac rhythm in patients with OHCA did not match the rhythm documented by EMS, we defined this as “discordant.” For example, if a patient with OHCA was noted to have an initial shockable rhythm by EMS, and the hospital team noted the initial hospital rhythm to be non-shockable (e.g. asystole or pulseless electrical activity), then we defined this as discordant documentation (**Figure 1**).

### Statistical Analysis

Descriptive statistics included the comparison of baseline characteristics between patients whose initial OHCA rhythm was documented by emergency medical services as shockable or non-shockable and by the hospital team as shockable or non-shockable. For outcome analyses, we examined concordance and discordance between EMS and in-hospital documentation of initial OHCA rhythm and the subsequent association with cardiac procedure incidence by calculating relative risk ratios. To investigate the impact of the initial ECG on hospital arrival, we computed the relative risk ratios for patients who received left or right heart catheterization, depending on whether they had ischemic changes on the hospital arrival ECG. We analyzed the data for both concordant and discordant documentation. Data analyses were performed with the use of RStudio version 4.2.1 (RStudio: Integrated Development for R. RStudio, PBC, Boston, MA URL http://www.rstudio.com/).

## RESULTS

### Baseline Characteristics

Out of 1051 CARES patients, 798 (75.9%) were associated and cross-referenced with hospital patient records. Within 24 hours of hospital arrival, 631 patients expired, leaving 167 patients for analysis. Of these 167 patients only 3 of these patients did not have hospital documentation about the initial OHCA rhythm, leaving 164 for final analysis (20.6% of the cross-referenced registry) for analysis (**Figure 1**). Baseline characteristics for these patients are displayed in **Table 1**. These patients had a mean age of 62.7 years. Of these 164 patients, 73 (44.5%) were female and 123 (75.0%) were African American. The utilization of an automated external defibrillator and a mechanical CPR support device as per EMS entry into the CARES registry are listed in **Table 3**.

**Table 1.**
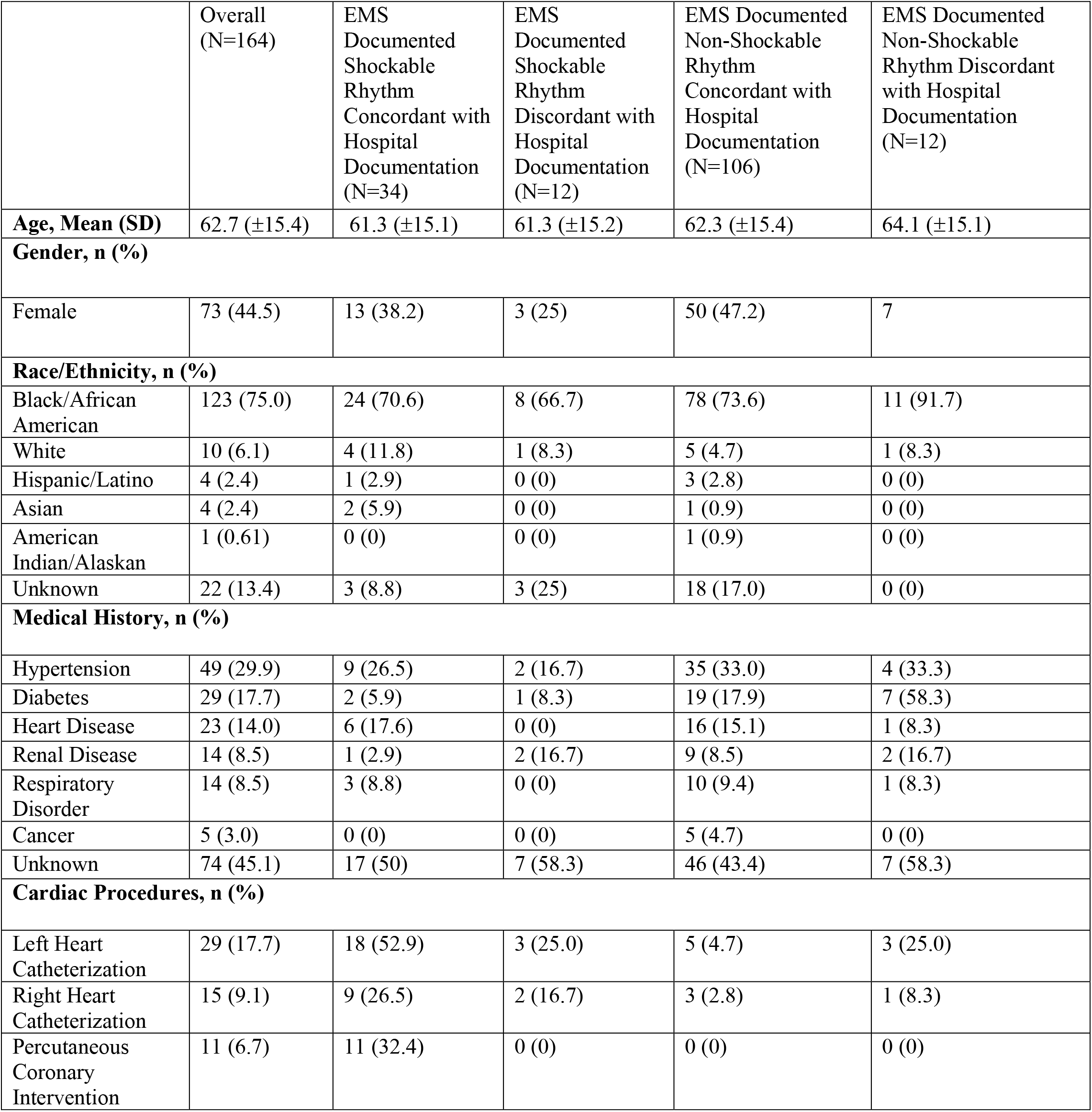
Baseline characteristics of overall cohort.

|  | Overall<br>(N=164) | EMS<br>Documented<br>Shockable<br>Rhythm<br>Concordant with<br>Hospital<br>Documentation<br>(N=34) | EMS<br>Documented<br>Shockable<br>Rhythm<br>Discordant with<br>Hospital<br>Documentation<br>(N=12) | EMS Documented<br>Non-Shockable<br>Rhythm<br>Concordant with<br>Hospital<br>Documentation<br>(N=106) | EMS Documented<br>Non-Shockable<br>Rhythm Discordant<br>with Hospital<br>Documentation<br>(N=12) |
| --- | --- | --- | --- | --- | --- |
| **Age, Mean (SD)** | 62.7 (±15.4) | 61.3 (±15.1) | 61.3 (±15.2) | 62.3 (±15.4) | 64.1 (±15.1) |
| **Gender, n (%)** |  |  |  |  |  |
| Female | 73 (44.5) | 13 (38.2) | 3 (25) | 50 (47.2) | 7 |
| **Race/Ethnicity, n (%)** |  |  |  |  |  |
| Black/African American | 123 (75.0) | 24 (70.6) | 8 (66.7) | 78 (73.6) | 11 (91.7) |
| White | 10 (6.1) | 4 (11.8) | 1 (8.3) | 5 (4.7) | 1 (8.3) |
| Hispanic/Latino | 4 (2.4) | 1 (2.9) | 0 (0) | 3 (2.8) | 0 (0) |
| Asian | 4 (2.4) | 2 (5.9) | 0 (0) | 1 (0.9) | 0 (0) |
| American Indian/Alaskan | 1 (0.61) | 0 (0) | 0 (0) | 1 (0.9) | 0 (0) |
| Unknown | 22 (13.4) | 3 (8.8) | 3 (25) | 18 (17.0) | 0 (0) |
| **Medical History, n (%)** |  |  |  |  |  |
| Hypertension | 49 (29.9) | 9 (26.5) | 2 (16.7) | 35 (33.0) | 4 (33.3) |
| Diabetes | 29 (17.7) | 2 (5.9) | 1 (8.3) | 19 (17.9) | 7 (58.3) |
| Heart Disease | 23 (14.0) | 6 (17.6) | 0 (0) | 16 (15.1) | 1 (8.3) |
| Renal Disease | 14 (8.5) | 1 (2.9) | 2 (16.7) | 9 (8.5) | 2 (16.7) |
| Respiratory Disorder | 14 (8.5) | 3 (8.8) | 0 (0) | 10 (9.4) | 1 (8.3) |
| Cancer | 5 (3.0) | 0 (0) | 0 (0) | 5 (4.7) | 0 (0) |
| Unknown | 74 (45.1) | 17 (50) | 7 (58.3) | 46 (43.4) | 7 (58.3) |
| **Cardiac Procedures, n (%)** |  |  |  |  |  |
| Left Heart Catheterization | 29 (17.7) | 18 (52.9) | 3 (25.0) | 5 (4.7) | 3 (25.0) |
| Right Heart Catheterization | 15 (9.1) | 9 (26.5) | 2 (16.7) | 3 (2.8) | 1 (8.3) |
| Percutaneous Coronary Intervention | 11 (6.7) | 11 (32.4) | 0 (0) | 0 (0) | 0 (0) |

### Baseline Characteristics and Outcomes by Initially Documented EMS Rhythm

Out of 164 patients included in our analysis, 46 (28.0%) patients’ initial cardiac rhythm was documented as shockable by EMS, while 118 (72.0%) patients’ initial cardiac rhythm was documented as non-shockable by EMS. Baseline characteristics for these patients are shown in **Table 2**. Of those patients whose initial cardiac rhythm was documented as shockable by EMS, 21 (45.7%) patients underwent left heart catheterization, and 11 (23.9%) patients underwent right heart catheterization. Of patients whose initial cardiac rhythm was documented as non-shockable by EMS, 8 (6.8%) patients underwent left heart catheterization, and 4 (3.4%) patients underwent right heart catheterization.

**Table 2.**
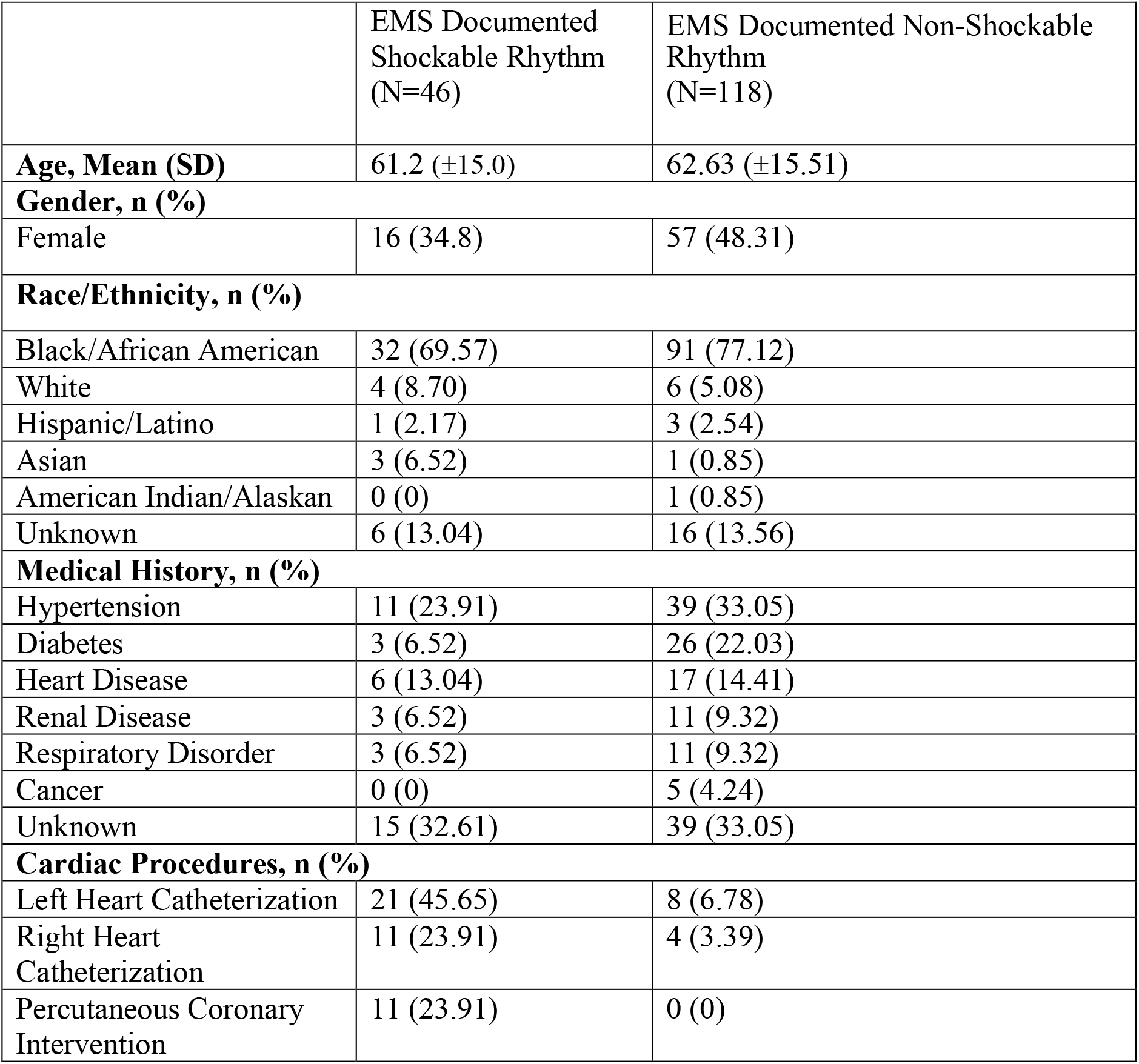
Baseline characteristics by initially documented emergency medical services (EMS) rhythm.

|  | EMS Documented Shockable Rhythm (N=46) | EMS Documented Non-Shockable Rhythm (N=118) |
| --- | --- | --- |
| **Age, Mean (SD)** | 61.2 (±15.0) | 62.63 (±15.51) |
| **Gender, n (%)** |  |  |
| Female | 16 (34.8) | 57 (48.31) |
| **Race/Ethnicity, n (%)** |  |  |
| Black/African American | 32 (69.57) | 91 (77.12) |
| White | 4 (8.70) | 6 (5.08) |
| Hispanic/Latino | 1 (2.17) | 3 (2.54) |
| Asian | 3 (6.52) | 1 (0.85) |
| American Indian/Alaskan | 0 (0) | 1 (0.85) |
| Unknown | 6 (13.04) | 16 (13.56) |
| **Medical History, n (%)** |  |  |
| Hypertension | 11 (23.91) | 39 (33.05) |
| Diabetes | 3 (6.52) | 26 (22.03) |
| Heart Disease | 6 (13.04) | 17 (14.41) |
| Renal Disease | 3 (6.52) | 11 (9.32) |
| Respiratory Disorder | 3 (6.52) | 11 (9.32) |
| Cancer | 0 (0) | 5 (4.24) |
| Unknown | 15 (32.61) | 39 (33.05) |
| **Cardiac Procedures, n (%)** |  |  |
| Left Heart Catheterization | 21 (45.65) | 8 (6.78) |
| Right Heart Catheterization | 11 (23.91) | 4 (3.39) |
| Percutaneous Coronary Intervention | 11 (23.91) | 0 (0) |

**Table 3.**
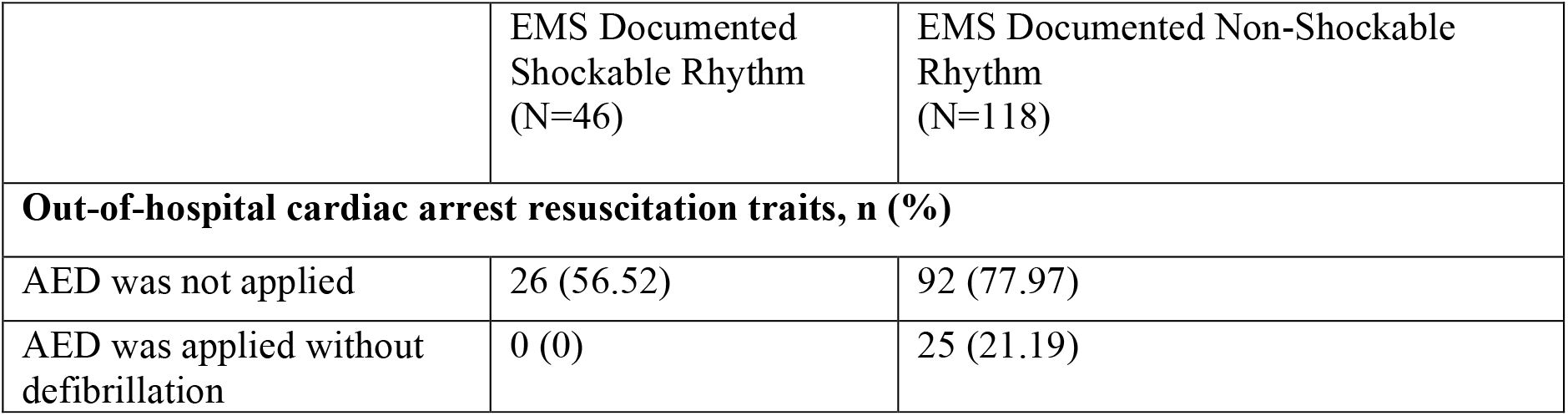
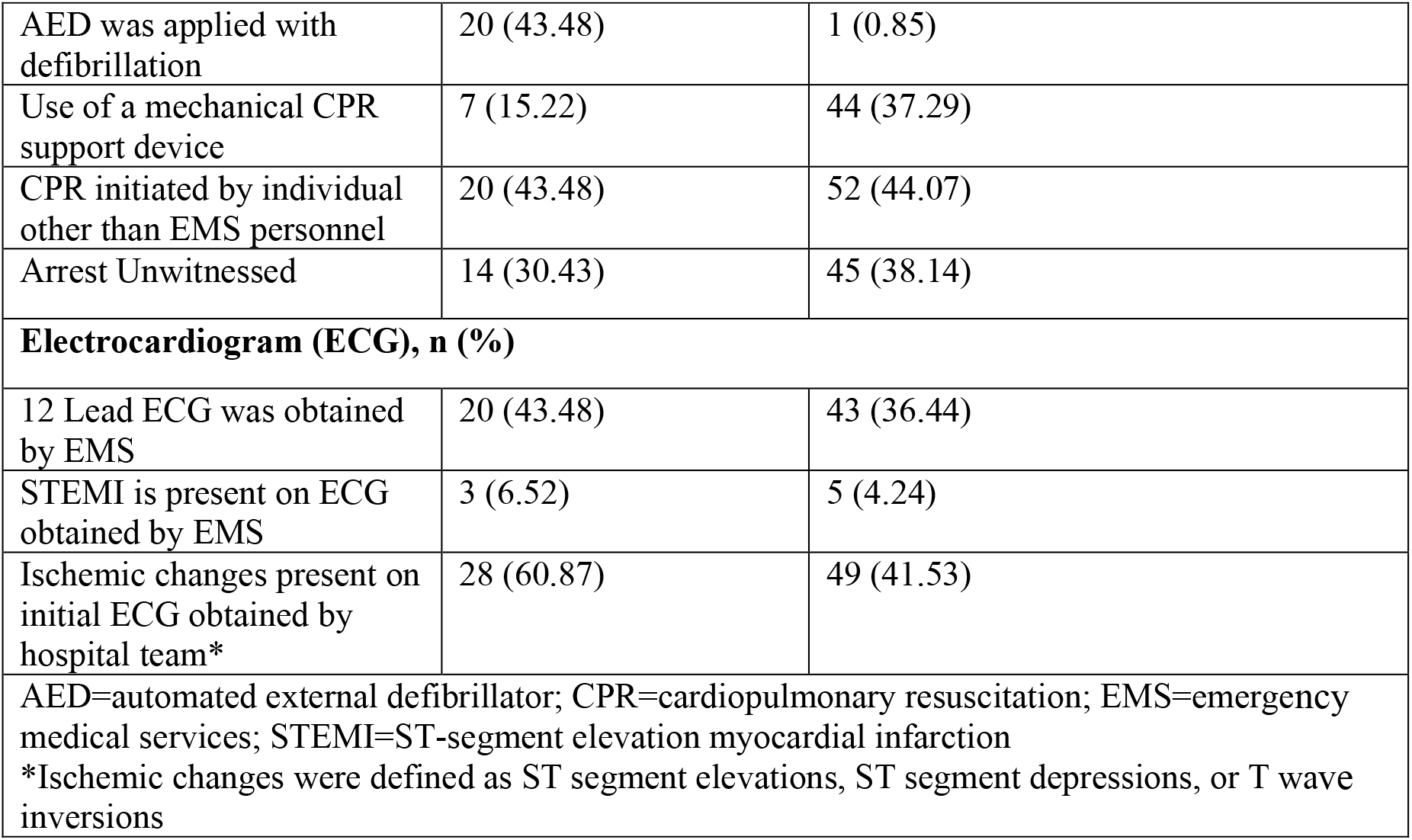
Out of hospital card iac arre st resu scit atio n characteristics by initially documented emergency medical services (EMS) rhythm.

### Influence of Concordance and Discordance on Outcomes by Initially Documented EMS Rhythm

The initial cardiac rhythm of 34 (20.7% of total analysis cohort) patients was concordantly documented as shockable by both EMS and the hospital team. However, for 12 (7.3% of total analysis cohort) patients, EMS documented the initial cardiac rhythm as shockable, while the hospital team documented the initial cardiac rhythm as non-shockable (**Table 1**). Thus, 26.1% of patients documented as having a shockable OHCA initial rhythm by EMS were discordantly documented as having a non-shockable OHCA initial rhythm in the inpatient documentation.

For patients whose initial cardiac rhythm was documented as shockable by EMS, the relative risk of left heart catheterization was 2.12 (95% RR CI: 0.76-5.93) when comparing hospital-documented shockable OHCA initial rhythm (i.e., “concordant”) versus hospital documented non-shockable OHCA initial rhythm (i.e., “discordant”) (**Figure 1**). Similarly, the relative risk of right heart catheterization was 1.59 (95% CI: 0.40-6.33) between the shockable OHCA concordant and discordant groups. (**Table 4**).

**Table 4.** Influence of concordance and discordance on relative risk of cardiac procedures by initially documented emergency medical services (EMS) rhythm.

|  | Initial EMS shockable rhythm comparing hospital concordance vs. discordance | Initial EMS non-shockable rhythm comparing hospital concordance vs. discordance |
| --- | --- | --- |
| Left Heart Catheterization Relative Risk Ratio (CI) | 2.12 (0.76-5.93) | 0.19 (0.05-0.69) |
| Right Heart Catheterization Relative Risk Ratio (CI) | 1.59 (0.40-6.33) | 0.34 (0.04-3.01) |

The initial cardiac rhythm of 106 (64.6% of total analysis cohort) patients was concordantly documented as non-shockable by both EMS and the hospital team. However, for 12 (7.3% of total analysis cohort) patients, EMS documented the initial cardiac rhythm as non-shockable, while the hospital team documented the initial cardiac rhythm as shockable (**Table 1**). Thus, 10.2% of patients documented as having a non-shockable OHCA initial rhythm by EMS were discordantly documented as having a shockable OHCA initial rhythm in the inpatient documentation.

For those patients whose initial cardiac rhythm was documented as non-shockable by EMS, the relative risk of left heart catheterization was 0.19 (95% CI: 0.05-0.69) when comparing hospital-documented non-shockable OHCA initial rhythm (i.e. “concordant”) and hospital documented shockable OHCA initial rhythm (i.e. “discordant”) (**Figure 1**). The relative risk of right heart catheterization was 0.34 (95% CI: 0.04-3.01) between the non-shockable OHCA concordant and discordant groups (**Table 4**).

Finally, patients with ischemic changes on their arrival hospital ECG had a 2.48 (95% CI: 1.20-5.12) times higher risk of undergoing left heart catheterization compared to those without such changes. These findings are summarized in **Table 5**.

**Table 5.**
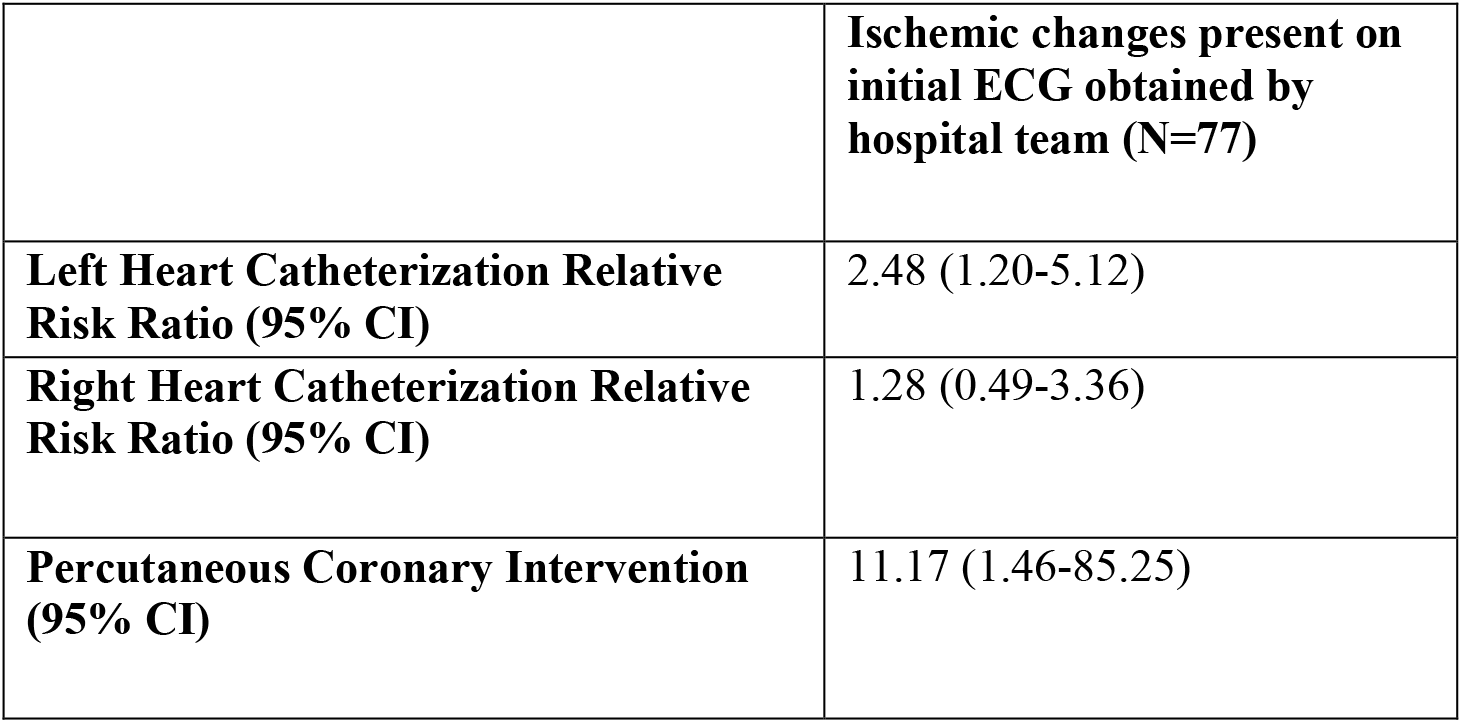
Influence of concordance and discordance on relative risk of cardiac procedures by presence of ischemic changes on initial ECG obtained by the hospital team, regardless of initial documented EMS rhythm.

## DISCUSSION

To our knowledge, we report the first analysis of the frequency and potential impact of discrepancies regarding the documentation of OHCA initial cardiac rhythm between EMS and the hospital team. In our analysis, 14.6% of OHCA patients surviving at least 24 hours after hospital presentation had discordance in the documentation of the initial cardiac arrest rhythm. While the reasons for the documentation discrepancy cannot be determined from our analysis, the presence of any communication gap between medical providers in the chain of survival may affect patient management and represents an opportunity for future work. We identified a potential association between initial cardiac rhythm discordance between EMS and hospital documentation with the relative frequency of invasive cardiac procedures after OHCA. The frequency of LHC was highest when there was agreement of a shockable rhythm and lowest when there was agreement of a non-shockable rhythm (52.9% and 4.7%, respectively). The frequency of LHC when there was hospital discordance in rhythm determination in cases of EMS determined shockable and non-shockable rhythm was numerically intermediate (25% and 25%, respectively) between the concordant results. In our primary analysis, these differences were only statistically significant for the EMS determined non-shockable rhythm patients. In both the EMS shockable and non-shockable OHCA cohorts, hospital documentation of a shockable OHCA initial rhythm was associated with a numerically higher relative risk of LHC. Such an association is clinically plausible as the inpatient clinical management after OHCA often relies on the initial cardiac rhythm.

Over the past several years survival rates for patients who experience OHCA have improved.[7] This progress in OHCA survival is multifactorial, but may be attributed to a combination of improvement in cardiopulmonary resuscitation (CPR) quality, early recognition of cardiac arrest, and the use of automated external defibrillators.[8],[9] Despite improvement in outcomes for patients who sustain OHCA, mortality remains high especially those who are reported to have non-shockable rhythms, with survival rates as low as 7%.[10],[11] Various hypotheses have been proposed for the morbidity and mortality associated with OHCA.

There are important regional variations in outcomes for patients with OHCA, [12],[13] in part attributed to care practice differences both during the pre-hospital and in-hospital phases of management.[12] The rate of rhythm disagreement in our study (14.6%) was noticeably higher compared to a population from the North Holland province of the Netherlands studied by Homma et al (4%), likely predominantly due to regional differences in first responder and EMS systems.[15], In the Homma et al study, EMS paramedics routinely send manual defibrillator ECG tracings to the study centers or, instead, when an AED was used the study personnel directly sent AED information to hospital teams if a shockable rhythm was detected.[16] This alerted physicians in that study region to the importance of AED information for patient outcomes in cases of OHCA. Neither of these robust data transmission practices exist for OHCA response in the Washington D.C. metropolitan population we studied. Additionally, regarding the pre-hospital setting, poorly economically resourced and more rural areas have been associated with worse outcomes in patients with OHCA.[14]

Regarding the in-hospital management regional variation, decision to proceed with coronary angiography is influenced by time from collapse to return of spontaneous circulation (ROSC) and other such predictive features for neurological recovery at some centers and not others.[6] Within medical centers, different specialists including anesthesiologists, cardiologists, emergency physicians, and intensivists may coordinate care of these patients leading to significant practice heterogeneity.[12] In addition to these factors, other areas of inconsistency such as the accuracy of documentation need to be identified and improved to further enhance outcomes.[17],[18] A solution to consider is electronically transferring the OHCA ECG directly to the hospital, making the prehospital ECG available in the patient’s electronic health records. Our report showed that there is important variation in the initial cardiac rhythm documentation, which emphasizes the need for accurate documentation and clear communication among different medical providers.

Improvements in communication practices during the transition of care from EMS to hospital teams, and among different hospital teams, is essential for better management of patients with OHCA.

## LIMITATIONS

There are numerous limitations to our study. Our cohort is both small and from a single institution, limiting the external validity of our findings. While the relative risk of documentation discrepancy on LHC incidence was significant for the EMS non-shockable rhythm cohort, the relative risk was not significant for the EMS shockable rhythm cohort. This difference may be due to small sample size given the wide confidence intervals in the smaller shockable rhythm cohort. Additionally, we were unable to associate all local OHCA patients from the CARES registry with hospital records using billing codes, and thus cannot exclude a significant impact of selection bias in our analysis cohort. However, if we were to assume that all 253 unmatched CARES registry patients either had concordant or discordant initial cardiac rhythm documentation, respectively, and had a similar rate of initial survival after hospital presentation (20.9%), the potential range for documentation concordance would be between 88.9% and 64.5% instead of the 85.46% in our analysis. Despite such exclusions, the demonstrated discrepancies suggest that discordance between EMS and hospital documentation remains prevalent. In addition, while the presence versus absence of ischemic ECG changes on initial hospital arrival ECG did associate with receiving left heart catheterization, the degree of the association was similar to initial rhythm concordance versus discordance. Another important limitation of our analysis is that we cannot independently verify the prehospital rhythm determination as pre-hospital rhythm strips were unavailable for review in our electronic medical record. However, available AED utilization data does support EMS documentation accuracy as only 1 (0.85%) patient in the EMS-documented non-shockable rhythm group with an AED applied received defibrillation and, similarly, all patients in the EMS-documented shockable rhythm group with an AED applied received defibrillation (Table 3).

Lastly, we acknowledge that various communities have varying approaches to and available resources for handling OHCA. Our study was conducted specifically focusing on individuals in our local community who had ready access to coronary angiography as determined to be clinically indicated.

## CONCLUSION

Discrepancies between EMS and hospital documentation regarding initial rhythm in out of hospital cardiac arrest is common. Such discrepancies may be associated with the relative risk of performance of invasive cardiac procedures for patients after OHCA. Dedicated investigation is required to understand how to improve both communication and documentation by medical staff managing OHCA patients.

## Data Availability

The data underlying this article will be shared on reasonable request to the corresponding author

## Abbreviations

OHCA: Out of hospital cardiac arrest
CA: Coronary angiography
EMS: Emergency medical services
CARES: Cardiac Arrest Registry to Enhance Survival

## REFERENCES

[1] T. Väyrynen, M. Kuisma, T. Määttä, and J. Boyd, “Medical futility in asystolic out-of-hospital cardiac arrest,” Acta Anaesthesiologica Scandinavica, vol. 52, no. 1, pp. 81–87, 2008, doi: 10.1111/j.1399-6576.2007.01461.x.

[2] J. Engdahl, A. Bång, J. Lindqvist, and J. Herlitz, “Factors affecting short- and long-term prognosis among 1069 patients with out-of-hospital cardiac arrest and pulseless electrical activity,” Resuscitation, vol. 51, no. 1, pp. 17–25, Oct. 2001, doi: 10.1016/S0300-9572(01)00377-X.

[3] S. Desch et al., “Angiography after Out-of-Hospital Cardiac Arrest without ST-Segment Elevation,” New England Journal of Medicine, vol. 0, no. 0, p. null, Aug. 2021, doi: 10.1056/NEJMoa2101909.

[4] K. B. Kern, “Optimal Treatment of Patients Surviving Out-of-Hospital Cardiac Arrest,” JACC: Cardiovascular Interventions, vol. 5, no. 6, pp. 597–605, Jun. 2012, doi: 10.1016/j.jcin.2012.01.017.

[5] B. McNally, A. Stokes, A. Crouch, and A. L. Kellermann, “CARES: Cardiac Arrest Registry to Enhance Survival,” Annals of Emergency Medicine, vol. 54, no. 5, pp. 674-683.e2, Nov. 2009, doi: 10.1016/j.annemergmed.2009.03.018.

[6] A. Lotfi et al., “SCAI expert consensus statement on out of hospital cardiac arrest,” Catheterization and Cardiovascular Interventions, vol. 96, no. 4, pp. 844–861, 2020, doi: 10.1002/ccd.28990.

[7] S. Yan et al., “The global survival rate among adult out-of-hospital cardiac arrest patients who received cardiopulmonary resuscitation: a systematic review and meta-analysis,” Crit Care, vol. 24, no. 1, p. 61, Feb. 2020, doi: 10.1186/s13054-020-2773-2.

[8] “Hüpfl: Chest-compression-only versus standard cardiopulm… - Google Scholar.” https://scholar.google.com/scholar_lookup?title=Chest-compression-only%20versus%20standard%20cardiopulmonary%20resuscitation%3A%20a%20meta-analysis&journal=Lancet&doi=10.1016%2FS0140-6736%2810%2961454-7&volume=376&issue=9752&pages=1552-1557&publication_year=2010&author=Hupfl%2CM&author=Selig%2CHF&author=Nagele%2CP (accessed Jan. 14, 2023).

[9] “Bystander Efforts and 1-Year Outcomes in Out-of-Hospital Cardiac Arrest | NEJM.” https://www.nejm.org/doi/full/10.1056/NEJMoa1601891 (accessed Jan. 14, 2023).

[10] E. Andrew, Z. Nehme, M. Lijovic, S. Bernard, and K. Smith, “Outcomes following out-of-hospital cardiac arrest with an initial cardiac rhythm of asystole or pulseless electrical activity in Victoria, Australia,” Resuscitation, vol. 85, no. 11, pp. 1633–1639, Nov. 2014, doi: 10.1016/j.resuscitation.2014.07.015.

[11] S. Saarinen, A. Kämäräinen, T. Silfvast, A. Yli-Hankala, and I. Virkkunen, “Pulseless electrical activity and successful out-of-hospital resuscitation – long-term survival and quality of life: an observational cohort study,” Scand J Trauma Resusc Emerg Med, vol. 20, no. 1, p. 74, Oct. 2012, doi: 10.1186/1757-7241-20-74.

[12] A. Proclemer et al., “Current practice in out-of-hospital cardiac arrest management: a european heart rhythm association EP network survey,” EP Europace, vol. 14, no. 8, pp. 1195–1198, Aug. 2012, doi: 10.1093/europace/eus232.

[13] D. Zive et al., “Variation in out-of-hospital cardiac arrest resuscitation and transport practices in the Resuscitation Outcomes Consortium: ROC Epistry–Cardiac Arrest,” Resuscitation, vol. 82, no. 3, pp. 277–284, Mar. 2011, doi: 10.1016/j.resuscitation.2010.10.022.

[14] S. Masterson, C. Teljeur, J. Cullinan, A. W. Murphy, C. Deasy, and A. Vellinga, “Out-of-hospital cardiac arrest in the home: Can area characteristics identify at-risk communities in the Republic of Ireland?,” International Journal of Health Geographics, vol. 17, no. 1, p. 6, Feb. 2018, doi: 10.1186/s12942-018-0126-z.

[15] P. C. M. Homma et al., “Transfer of essential AED information to treating hospital (TREAT),” Resuscitation, vol. 149, pp. 47–52, Apr. 2020, doi: 10.1016/j.resuscitation.2020.01.033.

[16] R. A. Waalewijn, R. de Vos, and R. W. Koster, “Out-of-hospital cardiac arrests in Amsterdam and its surrounding areas: results from the Amsterdam resuscitation study (ARREST) in Utstein style.,” Resuscitation, vol. 38, no. 3, pp. 157–167, Sep. 1998, doi: 10.1016/S0300-9572(98)00102-6.

[17] C. Hauw-Berlemont et al., “Emergency vs Delayed Coronary Angiogram in Survivors of Out-of-Hospital Cardiac Arrest: Results of the Randomized, Multicentric EMERGE Trial,” JAMA Cardiology, vol. 7, no. 7, pp. 700–707, Jul. 2022, doi: 10.1001/jamacardio.2022.1416.

[18] K. B. Kern et al., “Randomized Pilot Clinical Trial of Early Coronary Angiography Versus No Early Coronary Angiography After Cardiac Arrest Without ST-Segment Elevation: The PEARL Study,” Circulation, vol. 142, no. 21, pp. 2002–2012, Nov. 2020, doi: 10.1161/CIRCULATIONAHA.120.049569.

